# The proportion of seizure onset zone contacts resected is not associated with outcome following SEEG-guided resective epilepsy surgery in children

**DOI:** 10.1101/2021.09.18.21263722

**Authors:** Mehdi Khan, Aswin Chari, Kiran Seunarine, Christin Eltze, Friederike Moeller, Felice D’Arco, Rachel Thornton, Krishna Das, Stewart Boyd, J Helen Cross, M Zubair Tahir, Martin M Tisdall

## Abstract

**Purpose:** Children undergoing stereoelectroencephalography (SEEG)-guided epilepsy surgery represent a complex cohort. We aimed to determine whether the proportion of putative seizure onset zone (SOZ) contacts resected associates with seizure outcome in a cohort of children undergoing SEEG-guided resective epilepsy surgery.

**Methods:** Patients who underwent SEEG-guided resective surgery over a six-year period were included. The proportion of SOZ contacts resected was determined by co-registration of pre- and post-operative imaging. Seizure outcomes were classified as seizure free (SF, Engel class I) or not seizure-free (NSF, Engel class II-IV) at last clinical follow-up.

**Results:** Of 94 patients undergoing SEEG, 29 underwent subsequent focal resection of whom 22 had sufficient imaging data to be included in the primary analysis (median age at surgery of 10 years, range 5-18). Fifteen (68.2%) were SF and 7 (31.8%) NSF at median follow-up of 19.5 months (range 12-46). On univariate analysis, histopathology, was the only significant factor associated with SF (p<0.05). The percentage of defined SOZ contacts resected ranged from 25-100% and was not associated with SF (p=0.89). In a binary logistic regression model, it was highly likely that histology was the only independent predictor of outcome, although the interpretation was limited by pseudo-complete separation of the data.

**Conclusion:** Histopathology is a significant predictor of surgical outcomes in children undergoing SEEG-guided resective epilepsy surgery. The percentage of SOZ contacts resected was not associated with SF. Factors such as spatial organisation of the epileptogenic zone, neurophysiological biomarkers and the prospective identification of pathological tissue may therefore play an important role.

## Introduction

For carefully selected children with drug-resistant focal epilepsy, resective surgery is an established treatment, with up to of 70% achieving seizure freedom (SF).^1^ To delineate the resective target, a careful pre-surgical evaluation must be carried out and, in select candidates, this can involve the use of intracranial electroencephalography (iEEG) including stereoelectroencephalography (SEEG).

In cases that proceed directly to resective surgery (without iEEG), it has been shown that factors including complete resection of the magnetic resonance imaging (MRI)-visible lesion and histopathological diagnosis are key determinants of seizure freedom.^2^ The factors determining SF following SEEG-guided resective surgery have not been extensively studied. Correct delineation and subsequent resection of the putative seizure onset zone (SOZ) are potentially important factors. In adults, there is evidence that other markers such as interictal high frequency oscillations (HFOs)^3^ and ictal phase-locked high gamma (PLHG)^4^ may be better markers than the putative SOZ contacts, although these have largely been in patients undergoing subdural grid and strip recordings. The main aims of this study were to (a) quantify the proportion of SEEG-defined putative SOZ contacts resected by co-registering pre- and post-operative imaging and (b) identify factors, including the proportion of these contact resected, associated with post-operative SF in paediatric patients undergoing SEEG-guided resective epilepsy surgery at a single centre.

## Materials and Methods

### Setting

This was a single-centre, retrospective, observational study. STROBE guidelines were adhered to throughout this study.^5^ The project was registered with the Great Ormond Street Hospital (GOSH) R&D Office (19BI26). As it involved only retrospective use of routinely collected clinical data, formal ethical approval was not required.

### Participants

Paediatric patients (aged ≤18 years) who underwent SEEG at GOSH between 2014 and 2020 and subsequent resective epilepsy surgery were eligible for inclusion. Patients who had undergone previous epilepsy surgery, patients with tuberous sclerosis and patients with large structural abnormalities on MRI or computerised tomography (CT) imaging that would affect robust co-registration were excluded. As previously published, patients are selected for SEEG and subsequent surgical treatment based on a multidisciplinary team decision.^6^ This cohort overlaps partially with this previously published cohort and the technical details of the SEEG procedure and the clinical workflow are outlined elsewhere.^6–8^

### Data collection

Demographic, pre-surgical evaluation and SEEG variables were collected from electronic patient notes via a piloted proforma. Seizure onset patterns (SOP), which have been known to associate with post-surgical seizure outcome, were classified according to the methodology by Lagarde et al.^9^ Better prognosis has been reported with the presence of low voltage fast activity (LVFA) and for statistical analysis, the 8 patterns were dichotomised based on either the presence or absence of LVFA on SEEG.^9^ The main outcome measure was the Engel classification at last follow-up, dichotomised into SF (Engel class I) and not-seizure free (NSF, Engel classes II-IV). Assessment within 30 days of one year were classified as sufficient for one year follow-up. The SOZ contacts were taken as the ictal onset contacts as defined by the consultant neurophysiologist in the formal SEEG report.

### Segmentation & image registration

Individual electrode contact points from SEEG electrodes, localised using a CT scan and identified using SEEG assistant,^8^ were assigned voxel spaces and registered to the pre-operative imaging using *reg_aladin* (http://cmictig.cs.ucl.ac.uk/wiki/index.php/Reg_aladin). Post-operative volumetric MRI scans were used to manually delineate the area of resection using ITK-SNAP (v.3.X) and registered to the pre-operative imaging using *ANTS* (https://antspy.readthedocs.io/en/latest/index.html); the resection volume was excluded during this procedure to minimise distortion and account for brain collapse into the resection cavity, a common event following brain resections (Figure 1).^10–12^ The resected contacts were subsequently manually identified using *FSLeyes* to determine electrode overlap with the resection volume (Figure 1).

**Figure 1.**
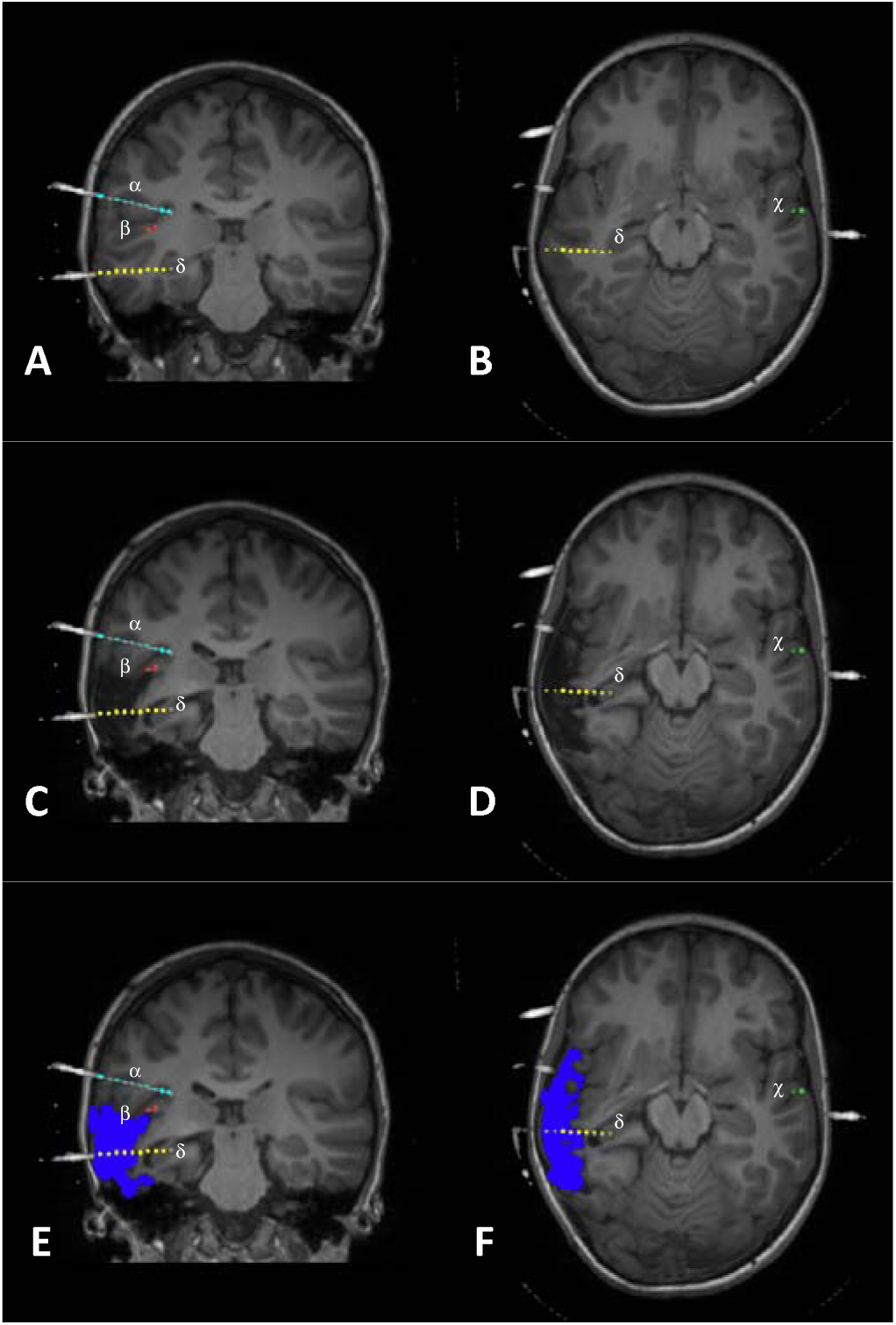
Co-registration of pre- and post-operative magnetic resonance images with SEEG electrodes’ computerised tomography images and segmentation images. MRI reconstructions of the resected seizure onset zone (SOZ) based on SEEG recordings, with co-registered coronal (left hand side panels) and axial (right hand side panels) scans. Four separate depth electrodes can be seen: electrode α targeting the right parietal operculum (light blue), electrode β targeting the right posterior superior temporal gyrus (STG) (red), electrode χ targeting the left STG (green) and electrode δ yellow targeting the right posterior hippocampus (yellow). Each electrode contact point is represented by a circle. Starting from the deepest one, contacts for each electrode are labelled numerically. In this example, pre-operative SEEG ictal recordings revealed a putative SOZ in the right STG (particularly the temporal operculum), represented by contacts δ[6, 7, 8, 9, 10] (**A and B**). These contacts are subsequently resected, as indicated in the post-operative scans (**C and D**). The resection area is then manually delineated to calculate the proportion of SOZ contacts resected, with the resection cavity marked in dark blue (**E and F**). In this case, 50% of the SEEG-defined SOZ contacts have been resected (note that other electrodes in the SEEG-defined, putative SOZ cannot be seen in these MRI planes. These MRI planes depicting 4 electrodes have only been chosen for representation purposes).

### Statistical analysis

Appropriate statistical tests (chi-sq for categorical variables & Mann-Whitney U-test for continuous variables) were used to assess association between variables of interest and seizure freedom. A post-hoc, residual analysis was performed to assess individual sub-categorical significance if predictors with more than two categories were significant, using Bonferroni-adjusted p-values to determine significance.^13^ A binary logistic regression model was fitted to assess for independent predictors for the post-operative Engel outcome.

Patients without sufficient imaging (either pre-operative or post-operative) were excluded from the primary analysis but were included in a sensitivity analysis (identical to the primary analysis but excluding variables determined by image analysis) to determine the robustness of the primary analysis.

All statistical tests were performed on SPSS v27. Images were created using GraphPad Prism 9.1. Statistical significance was taken at p-values < 0.05.

## Results

Between 2014-2020, 94 patients underwent a total of 98 SEEG explorations and 29 underwent subsequent resective surgery. Twenty-two patients had sufficient imaging data for the primary analysis (Table 1) and all 29 were included in the subsequent sensitivity analysis (Supplementary Table 1).

**Table 1:**
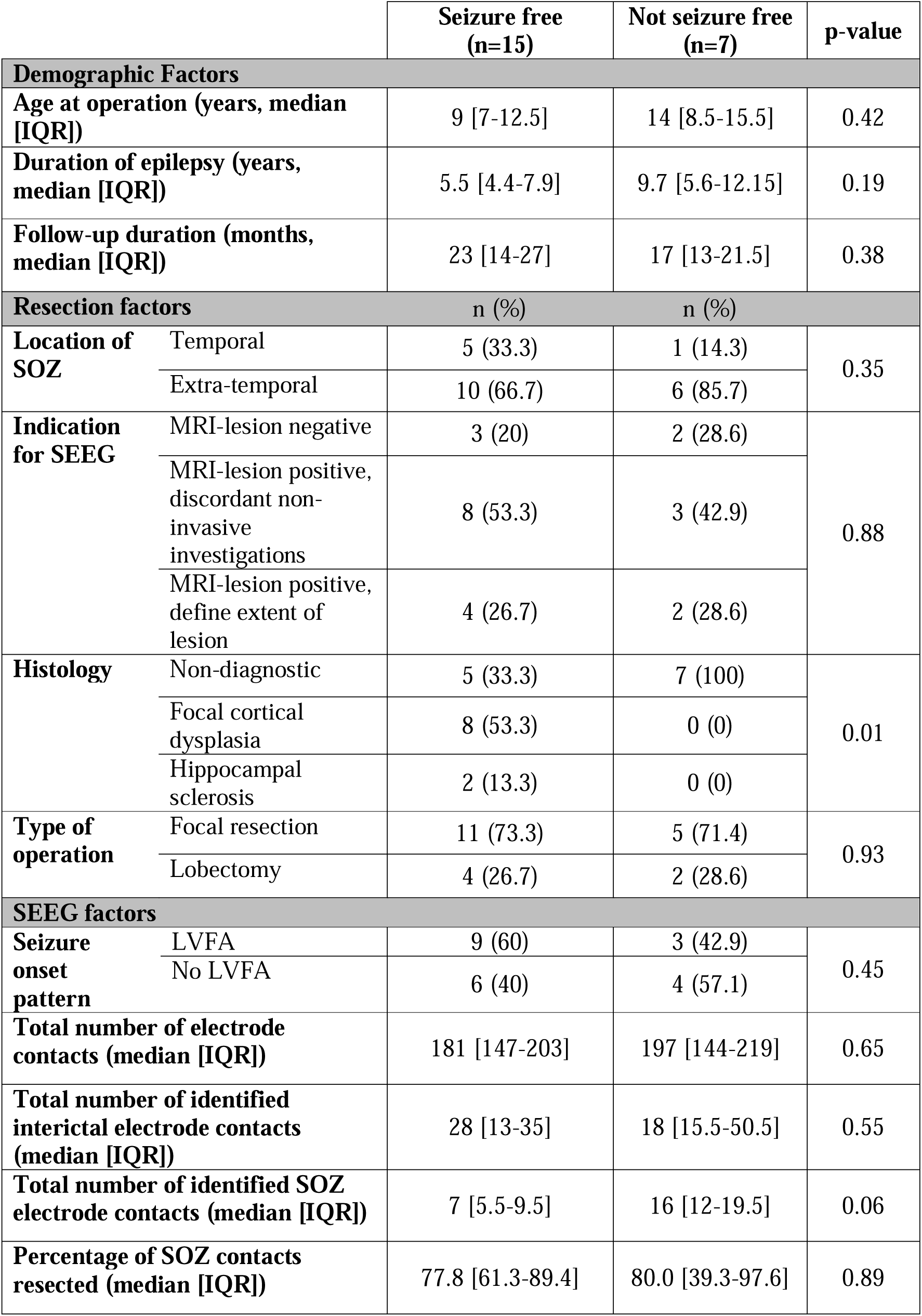
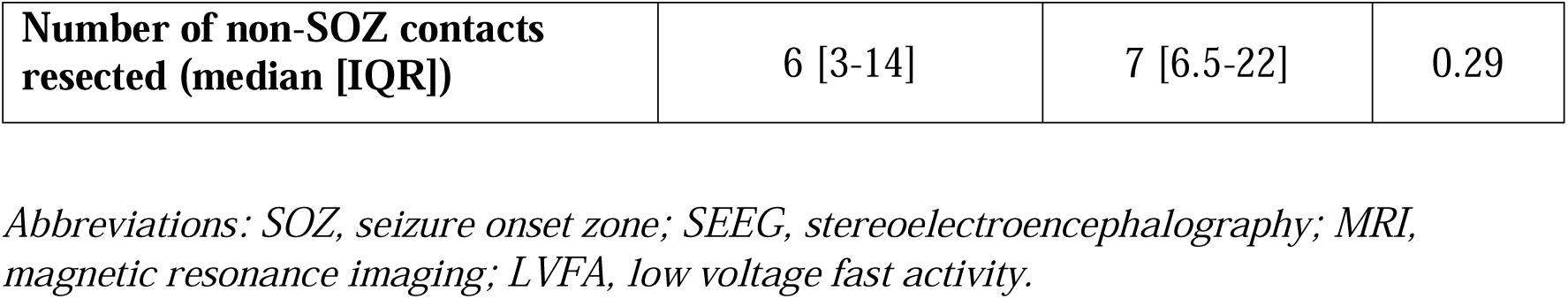
Results of univariate statistical comparisons during primary analysis for differences between patients that were seizure free and not seizure free.

|  |  | Seizure free<br>(n=15) | Not seizure free<br>(n=7) | p-value |
| --- | --- | --- | --- | --- |
| Demographic Factors |  |  |  |  |
| Age at operation (years, median [IQR]) |  | 9 [7-12.5] | 14 [8.5-15.5] | 0.42 |
| Duration of epilepsy (years, median [IQR]) |  | 5.5 [4.4-7.9] | 9.7 [5.6-12.15] | 0.19 |
| Follow-up duration (months, median [IQR]) |  | 23 [14-27] | 17 [13-21.5] | 0.38 |
| Resection factors |  | n (%) | n (%) |  |
| Location of SOZ | Temporal | 5 (33.3) | 1 (14.3) | 0.35 |
|  | Extra-temporal | 10 (66.7) | 6 (85.7) |  |
| Indication for SEEG | MRI-lesion negative | 3 (20) | 2 (28.6) | 0.88 |
|  | MRI-lesion positive, discordant non-invasive investigations | 8 (53.3) | 3 (42.9) |  |
|  | MRI-lesion positive, define extent of lesion | 4 (26.7) | 2 (28.6) |  |
| Histology | Non-diagnostic | 5 (33.3) | 7 (100) | 0.01 |
|  | Focal cortical dysplasia | 8 (53.3) | 0 (0) |  |
|  | Hippocampal sclerosis | 2 (13.3) | 0 (0) |  |
| Type of operation | Focal resection | 11 (73.3) | 5 (71.4) | 0.93 |
|  | Lobectomy | 4 (26.7) | 2 (28.6) |  |
| SEEG factors |  |  |  |  |
| Seizure onset pattern | LVFA | 9 (60) | 3 (42.9) | 0.45 |
|  | No LVFA | 6 (40) | 4 (57.1) |  |
| Total number of electrode contacts (median [IQR]) |  | 181 [147-203] | 197 [144-219] | 0.65 |
| Total number of identified interictal electrode contacts (median [IQR]) |  | 28 [13-35] | 18 [15.5-50.5] | 0.55 |
| Total number of identified SOZ electrode contacts (median [IQR]) |  | 7 [5.5-9.5] | 16 [12-19.5] | 0.06 |
| Percentage of SOZ contacts resected (median [IQR]) |  | 77.8 [61.3-89.4] | 80.0 [39.3-97.6] | 0.89 |

|  |  |  |  |
| --- | --- | --- | --- |
| **Number of non-SOZ contacts resected (median [IQR])** | 6 [3-14] | 7 [6.5-22] | 0.29 |

*Abbreviations: SOZ, seizure onset zone; SEEG, stereoelectroencephalography; MRI, magnetic resonance imaging; LVFA, low voltage fast activity.*

### Primary Analysis

The median duration of epilepsy of these 22 patients was 6.3 years (range 2.5-14.5). At a median age of 10 years (range 5-18), 16 (72.7%) patients underwent a focal resection and 6 (27.3%) underwent a larger lobar resection (lobectomy), respectively. The indications for SEEG exploration varied, with 11 (50%) patients classified as ‘MRI-lesion positive, discordant non-invasive investigations’, 6 (27.3%) as ‘MRI-lesion positive, define extent’ and 5 as ‘MRI-lesion negative’. The seizure onset pattern included LFVA in 12 (54.5%) and did not in 10 (45.5%). On histopathological examination, 12 (54.5%) were non-diagnostic, 8 (36.4%) were focal cortical dysplasia (FCD) (2 FCD type 2a, 5 FCD type 2b and 1 FCD type 2 not otherwise specified), and 2 (9.1%) were hippocampal sclerosis (HS).

Overall, 15 (68.2%) patients were SF, and 7 (31.8%) were NSF at median follow-up of 19.5 months (range 12-46 months); all patients had 1 year follow-up at minimum. There were no operative complications or new neurological deficits from the resective operations. Histopathological diagnosis was the only significant factor associated with seizure outcome (p<0.05) (Table 1, Figure 2a); on post-hoc analysis, FCD associated with increased likelihood of SF (100% SF, corrected p-value = 0.04) and ND was associated with decreased likelihood of SF (45.5% SF, corrected p-value = 0.009). All other pre-surgical and SEEG factors did not significantly associate with seizure outcome (Table 1). The percentage of SOZ contacts resected did not significantly associate with seizure outcome (p=0.89), although there was a trend that number of SOZ contacts was lower in the SF group (Figure 2b-c).

**Figure 2:**
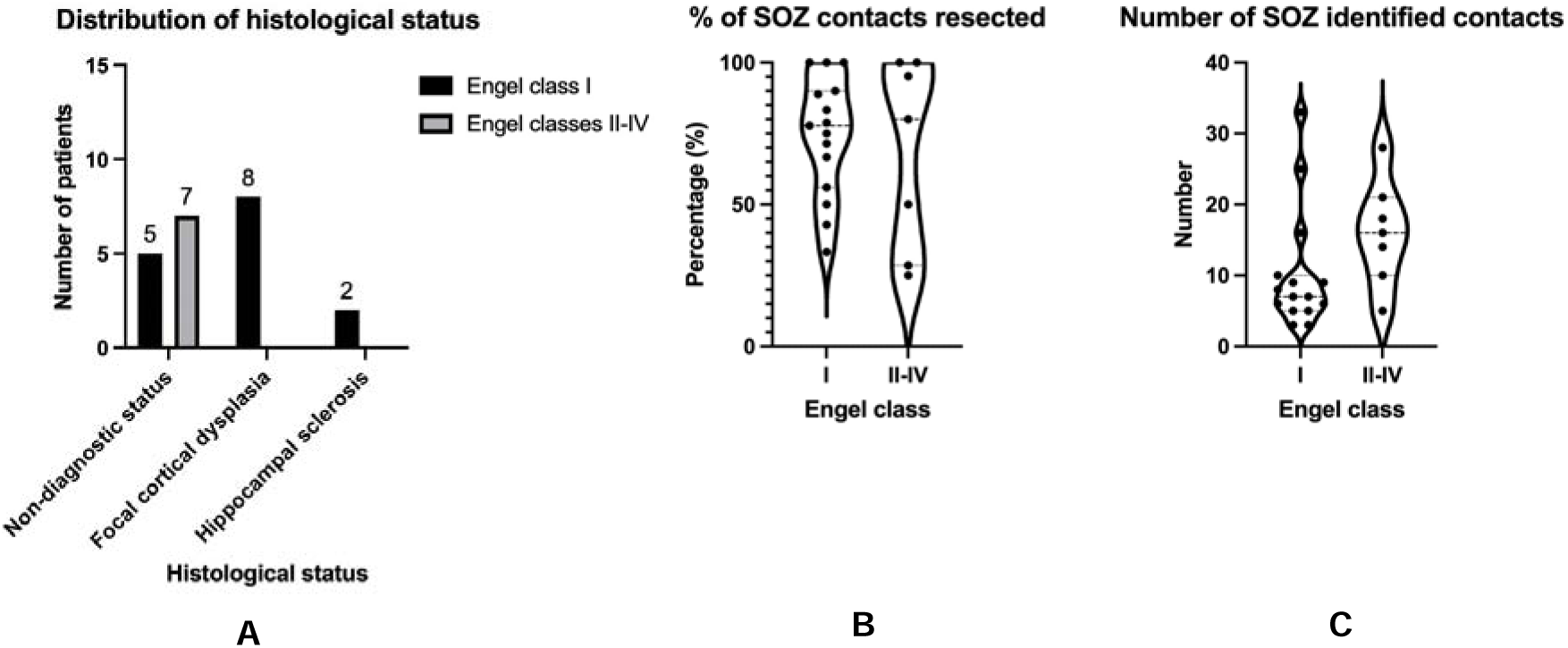
Association of pertinent variables with post-operative seizure outcome. (A) Histological status statistically associated with outcome (p=0.01). (B) Percentage of seizure onset zone contacts resected was not significantly associated with outcome (p=0.89). (C) Although not statistically significant, there were fewer SOZ contacts identified in the seizure-free cohort (median 7) compared to the non-seizure free cohort (median 16, p=0.06).

A binary logistic regression model with backwards elimination was fitted to ascertain the effects of pre-operative & SEEG factors on post-operative seizure outcome. Variables with a p≤0.3 on univariate analysis were selected, resulting in a statistically significant model (p<0.05). The model explained 56% of the variance in seizure outcome and correctly classified 77.3% of cases. The model parameter estimates failed to converge due to pseudo-complete separation of the seizure outcome data with the histopathological classification as all the FCD & HS patients had a favourable outcome. Although this resulted in the p-value for histopathology being uninterpretable, it is highly likely that the variable was an independent significant predictor in the model.^14^ All other variables were not independent predictors of seizure outcome.

### Sensitivity Analysis

In a sensitivity analysis including a total of 29 patients, all the above univariate and binary logistic regression statistical outcomes were confirmed, indicating robust outcomes and no systematic bias in the patients that did/did not have adequate imaging (Supplementary Table 1).

## Discussion

With post-operative SF rates of up 70%,^1^ epilepsy surgery is an accepted treatment option for children with focal drug-resistant epilepsy. Numerous studies over the years have assessed predictors of post-operative SF in these patients. The importance of the extent of resection of the MRI-visible lesion is well known, with complete resection being a significant factor determining SF.^2^ However, comparatively little is known about the corollary of this ‘extent of resection’ in the context of SEEG-guided resective epilepsy surgery. In our series, we identify that high rate of seizure freedom (66% of the entire cohort) is possible following SEEG-guided resective surgery. We identified that the percentage of SEEG contacts resected did not associate statistically with SF both in univariate and binary logistic regression analyses. Histology was the only significant factor predicting seizure outcome, with FCD being associated with SF status and ND with NSF status.

These findings highlight the limitations of current neurophysiological paradigms in delineating the SOZ to guide resective surgery in children undergoing SEEG. The different seizure onset patterns were not associated with seizure outcome, which may be due to a difference in the sample size or a biological difference between adults and children undergoing intracranial evaluation. Interestingly, there was a trend to suggest that a lower number of SOZ contacts associated with seizure freedom suggesting that a more focal SOZ may be more favourable than widespread network onset, in agreement with the findings of Lagarde et al.^9^ A smaller number of electrode contacts indicates a compact, localised SOZ which is more likely to indicate a discrete focal brain abnormality.^15^

If the resection involves an FCD, seizure freedom is highly likely irrespective of the percentage of neurophysiologically-defined SOZ contacts resected whilst a ND histology is associated with less favourable outcomes. In these cases, it could be that the SEEG resulted in mis-localisation of the actual pathological abnormality (e.g., an FCD elsewhere) or there is no clear abnormality, both of which have been associated with poorer outcomes. Alternatively, it could be that there are alternative pathological entities that are newly being identified such as mild malformation of cortical development with oligodendroglial hyperplasia and epilepsy (MOGHE), associated with poorer outcomes.^16^

The results also highlight the difficulty of ensuring that all intended contacts are resected following SEEG. Whilst some of the variability may be down to registration error, resecting intended contacts may be limited by functional boundaries or geographically separated contacts which are not all amenable to being resected. Advances in intraoperative navigation (including adding the SEEG electrode targets) may aid achieving the intended resections.

The results also highlight the expressed need for novel computational analyses and perspectives on interpreting SEEG data.^15^ This has been acknowledged for a number of years and, recently, multiple novel approaches including identification of novel SOZ biomarkers (ictal high gamma activity and interictal HFOs),^3,4^ comparison with normalised connectivity atlases^17,18^ and novel network synchronisability metrics^19^ have been used to try and explain surgical failures although these computational analyses are, to our knowledge, not yet being used to guide routine clinical practice. Prospective multicentre evaluation of these technologies is crucial to prove efficacy prior to widespread adoption.

### Limitations

There are several limitations to this study. Despite robust statistical outcomes following sensitivity analyses, this study is retrospective and its generalisability to other centres is unknown. Additionally, a small sample size increases the likelihood of type II errors. The neurophysiological definition of SOZ based on the clinical report was not interrogated further and taken at face value.

## Conclusion

In this single centre study analysing 22 patients undergoing SEEG-guided resective epilepsy surgery, we found that the proportion of SEEG-defined SOZ contacts resected is not a significant predictor of post-operative seizure outcome. The histopathology was the only significant predictor of seizure outcome; all patients with a diagnosis of FCD were seizure-free at last follow-up. All other pre-operative, operative and post-operative factors did not significantly associate with seizure outcome.

## Data Availability

N/A

## Abbreviations

SF: Seizure freedom
iEEG: Intracranial electroencephalography
SEEG: Stereoelectroencephalography
MRI: Magnetic resonance imaging
SOZ: Seizure onset zone SF Seizure freedom
HFO: High frequency oscillations
PLHG: Phase-locked high gamma
CT: Computerised tomography
SOP: Seizure onset patterns
LVFA: Low voltage fast activity
NSF: Not-seizure free
FCD: Focal cortical dysplasia
HS: Hippocampal sclerosis

## Disclosures

AC is supported by a Great Ormond Street Hospital Children’s Charity Surgeon Scientist Fellowship. This work was supported by the National Institute of Health Research–Great Ormond Street Hospital Biomedical Research Centre.

## Notes

### Competing Interest Statement

The authors have declared no competing interest.

### Author Declarations

This retrospective study was conducted following approval from the Joint Research Office of Great Ormond Street Hospital & University College London Institute of Child Health (project ID 19BI26). As it was a retrospective study using routinely collected clinical data, formal ethical approval was waived by the Joint Research Office of Great Ormond Street Hospital & University College London Institute of Child Health.

