## Supplementary Table 1 for "The proportion of seizure onset zone contacts resected is not associated with outcome following SEEG-guided resective epilepsy surgery in children"

**Supplementary Table 1: Results of univariate statistical comparisons during sensitivity analysis for differences between patients that were seizure free and not seizure free.**

| | | Seizure free (n=19) | Not seizure free (n=10) | p-value |
|---|---|---|---|---|
| **Demographics factors** | | | | |
| **Age at operation (years, median [IQR])** | | 10 [7.5-16] | 17 [14.5-18] | 0.321 |
| **Duration of epilepsy (years, median [IQR])** | | 7 [4.65-8.4] | 9.85 [6.65-12.7] | 0.168 |
| **Follow-up duration (months, median [IQR])** | | 24.5 [13.8-27] | 17 [14-18] | 0.612 |
| **Resection factors** | | n (%) | n (%) | |
| **Location of SOZ** | Temporal | 8 (42.1) | 4 (40) | 0.913 |
| | Extra-temporal | 11 (57.9) | 6 (60) | |
| **Indication for SEEG** | MRI-lesion negative | 5 (26.3) | 5 (50) | 0.403 |
| | MRI-lesion positive, discordant non-invasive investigations | 10 (52.6) | 3 (30) | |
| | MRI-lesion positive, define extent of lesion | 4 (21.1) | 2 (20) | |
| **Histology** | Non-diagnostic | 6 (31.6) | 10 (100) | 0.002 |
| | Focal cortical dysplasia | 10 (52.6) | 0 (0) | |
| | Hippocampal sclerosis | 3 (15.8) | 0 (0) | |
| **Type of operation** | Focal resection | 13 (68.4) | 5 (50) | 0.331 |
| | Lobectomy | 6 (31.6) | 5 (50) | |
| **SEEG factors** | | | | |
| **Location of SOZ** | LVFA | 13 (68.4) | 4 (40) | 0.14 |
| | No LVFA | 6 (31.6) | 6 (60) | |
| **Total number of identified interictal electrode contacts (median [IQR])** | | 27 [13.5-31.8] | 16 [13-38] | 0.95 |

Abbreviations: SOZ, seizure onset zone; SEEG, stereoelectroencephalography; MRI, magnetic resonance imaging; LVFA, low voltage fast activity.
